# Activity-Induced Changes in Pain and Knee Range of Motion in Adults With Knee Osteoarthritis

**DOI:** 10.64898/2026.05.04.26352365

**Authors:** Julien A. Mihy, Mayumi Wagatsuma, Elisa S. Arch, Katie A. Butera, Stephen M. Cain, Jocelyn F. Hafer

## Abstract

**Background:** Pain with movement is common in adults with knee osteoarthritis (OA), but the effect of movement-evoked pain on gait is not well understood. This relationship is vital to understand as gait mechanics are associated with OA initiation and progression. Our current understanding of acute changes in pain and gait stems from extended bouts of walking, however these bouts likely don’t represent real-world behavior. Therefore, understanding how gait changes with shorter, more intense bouts of activity may provide valuable insight into the pain experience.

**Methods:** Adults with (n=19) and without (n=19) knee OA wore inertial measurement units (IMUs) while completing bouts of walking before and after two bouts of stair navigation (two flights). We tested whether pain and gait (speed, stride length, and lower extremity joint ranges of motion (ROM)) changed differently between adults with and without knee OA in response to multiple bouts of stair activity.

**Findings:** There were no significant interactions between group and stair bouts for any variable. When stratifying the OA group by those who did and did not experience pain, those who experienced a change in pain also had a greater change in early stance knee ROM in response to bouts of stairs.

**Interpretation:** The observed changes suggest that knee kinematics may be more sensitive to acute changes in pain than gait speed or stride length. These differences were detectable using IMUs and therefore our results support the use of IMUs to measure concurrent pain and gait mechanics in less controlled and real-world settings.

## 1. Introduction

Pain in response to movement is common in adults with knee osteoarthritis (OA), but the effect of movement-evoked pain on gait is not well understood. This limited understanding is in part due to previous study designs. Most studies aiming to understand the relationship between pain and gait mechanics assess how walking mechanics differ with disease severity (Costello et al., 2021; Heiden et al., 2009; Henriksen et al., 2012; Teichtahl et al., 2006), subjective pain questionnaire scores (Costello et al., 2021; Maly et al., 2008; O’Connell et al., 2016), or in response to pain relief (Schnitzer et al., 1993; Shrader et al., 2004). These studies typically focus on differences in knee adduction moments (KAM) or knee flexion moments as they approximate frontal and sagittal plane joint loads, respectively. Although we may assume that greater joint loads would be associated with greater pain during gait, a meta-analysis revealed only a medium positive relationship between peak KAM and pain in those with medial compartment knee OA (Hutchison et al., 2023). This finding reflects the inconsistencies across studies with some reporting a positive association between KAM and gait-related pain (Amin et al., 2004; Miyazaki et al., 2002; Thorp et al., 2007) and others reporting a negative association (Heiden et al., 2009; Henriksen et al., 2012; Schnitzer et al., 1993; Teichtahl et al., 2006). These inconsistencies demonstrate the need to further investigate the complex relationship between pain and gait via different methodological approaches.

The effect of pain on gait is vital to understand as gait mechanics are associated with OA initiation and progression. This relationship is difficult to assess because gait can both evoke pain or change in response to pain. Study designs that use bouts of activity to provoke pain are useful for differentiating between gait mechanisms that are protective and those that are in response to pain. To differentiate these mechanisms, a few studies have measured the dynamic relationship between changes in pain and gait in response to prolonged bouts of continuous walking (Boyer and Hafer, 2019; Farrokhi et al., 2017; Gustafson et al., 2019). These studies identified that 20-30 minutes of continuous treadmill walking can elicit increases in pain. Boyer and Hafer found that roughly 40% of participants with knee OA had an increase in pain in response to walking. Those who experienced an increase in pain had greater decreases in knee flexion and adduction moments compared to those with knee OA who did not experience a change in pain. Farrokhi et al. and Gustafson et al. (same cohort) used continuous and interval treadmill walking and identified no increase in pain with interval walking and increases in pain after 15-minutes of continuous walking. They used musculoskeletal modeling to estimate knee contact forces and found that 30 minutes of walking led to increased knee joint loading. While these studies provide insight into the influence of activity-induced pain on gait mechanics, their prolonged walking protocols may not resemble daily activity. Only 13-40% of adults with knee OA meet the minimum guidelines for moderate to vigorous physical activity, meaning that few individuals would be likely to complete 20 or 30 minute bouts of walking in daily life (Wallis et al., 2013). However, pain is a common daily symptom and therefore understanding how gait changes in response to common shorter, more intense bouts of activity may provide new insight into the pain experience and better represent real-world activity.

Stairs are a common activity of daily living that are often painful even in relatively short bouts and in early stages of knee OA (Hensor et al., 2015). Examining how gait and pain change in response to bouts of stairs may provide a more consistent and stronger pain response than in prior work and be more in line with an activity that patients complete in daily life. Multiple bouts of stair activity may influence gait mechanics independently of pain, therefore it is important to understand how adults with knee OA respond to such bouts of activity compared to their healthy counterparts. Therefore, the purpose of this study was to determine how pain and gait change in response to multiple bouts of stair activity in those with and without knee osteoarthritis using methods that could be translated to real-world settings (i.e., inertial measurement units [IMUs]). Because flexion ranges of motion (Boyer and Hafer, 2019; Maly et al., 2008) and gait speed (Bindawas, 2016) are associated with pain intensity and can be accurately measured with IMUs (Hafer et al., 2020), we hypothesized that adults with knee OA would have greater decreases in gait speed, stride length, and hip, knee, and ankle ranges of motion (ROM) during level gait compared to their healthy counterparts following bouts of stair activity. Our knee ROM measures included ROM across the gait cycle and early stance knee ROM because of its influence on joint loads during weight acceptance (Creaby et al., 2013).

## 2. Methods

### 2.1. Participants

A power analysis was conducted using G*Power version 3.1.9.7 (Faul et al., 2007) for sample size estimation, based on previous work finding a medium effect size (*d =* 0.42) when comparing gait speed before and after an injection to relieve pain in individuals with knee OA (Shrader et al., 2004). With stairs being a common source of pain in adults with knee OA, we expected a similar to greater effect size. Therefore, the minimum sample size needed to detect an interaction effect (greater changes in response to stairs in the adults with knee OA) with a medium effect size (f = 0.21) with significance criterion of α = 0.05 and power = 0.80, was N = 38 (19 per group).

The participants with knee OA met the American College of Rheumatology clinical criteria for knee OA by having the presence of occasional to frequent knee pain, minimal morning joint stiffness, and no palpable joint warmth. All participants were able to walk continuously for 30 minutes without assistive devices, and had no history of a major lower extremity surgery (arthroscopies >1 year prior were allowed), no current pain (besides the symptomatic knee), and no history of cardiovascular, pulmonary, or neurological conditions that limited daily activity. All participants completed a University of Delaware (Newark, DE, USA) IRB-approved informed consent before participating.

### 2.2. Data Collection

#### 2.2.1. Inertial Measurement Units

Five IMUs (Opal v2, APDM/Clario, Philadelphia, PA) collected 3-axis accelerometer, gyroscope, and magnetometer signals continuously at 128 Hz in synchronized logging mode. IMUs were placed by study personnel on the participants’ sacrum, thigh and shank of the right (healthy) or most symptomatic limb (OA), and bilateral feet. Thigh and shank sensors were placed on the lateral aspect of the segment’s midpoint and foot sensors were placed on the dorsum of the participants’ shoes (Figure 1).

**Figure 1.**
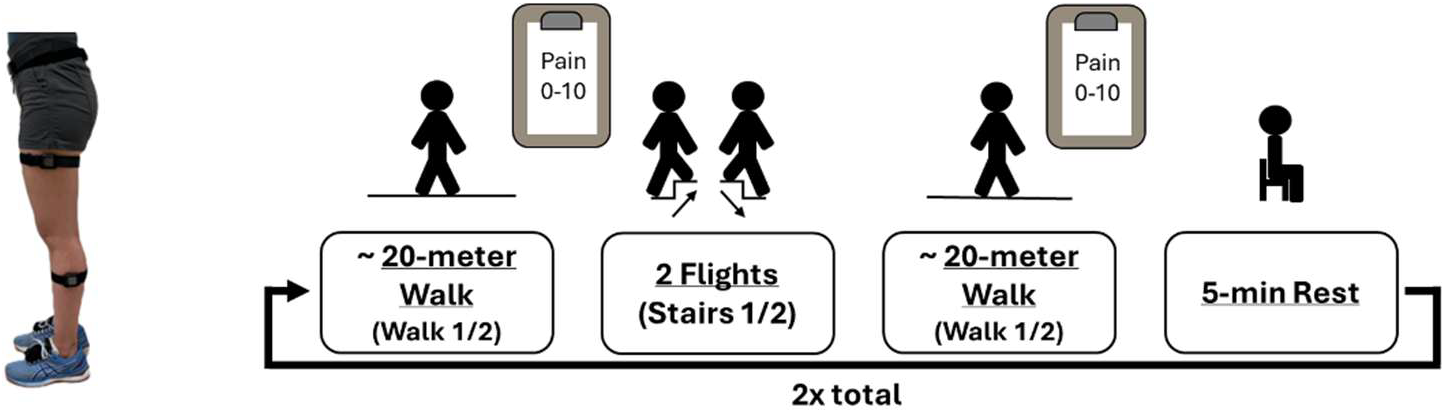
**Left:** Visualization of sensor placements. **Right:** Protocol schematic.

#### 2.2.2. Protocol

Data collection began with participants completing a series of questionnaires including the KOOS (Knee Injury and Osteoarthritis Outcome Survey). Following questionnaires, participants completed two clinical tasks (sit-to-stand and 40-m walk test). The KOOS and clinical tests were used for participant characterization. Participants were then fitted with the IMUs and completed functional movements and postures: quiet standing, toe touches, and a short bout of straight-line walking (Mihy et al., 2026).

Following the functional movements, participants were escorted to a nearby stairwell that was adjacent to a ∼20 m hallway. Participants then completed a short bout of walking down and back the hallway (Figure 1, Walk 1). Following the walk, they were asked to report the peak pain level experienced as they completed that walk. Participants then ascended and descended two flights of stairs (S1; 1 flight=25 stairs). After the stairs, participants immediately completed another walk down and back the hallway (Walk 2) and reported what their peak pain was during that walk. Following Walk 2, all participants took a 5-minute seated rest. Participants were allowed an additional 5 minutes if needed, but none required more rest. After resting, participants repeated the process with another walk (Walk 3), stair navigation (S2), and walk (Walk 4) totaling four walks of interest with their corresponding pain levels (Figure 1). Throughout data collection, researchers had an IMU used as a trigger to identify the times of the functional tasks and four walks of interest.

### 2.3. Data Processing

IMU data processing began by identifying the data used for the functional orientation procedures. The functional data included quiet standing, toe touches, and straight-line walking and were identified using trigger button pushes at the start and stop of each movement. Sensor data were oriented to functional (sensor-to-segment orientation) reference frames using quiet standing data and data from toe touches (sacrum) or straight-line walking (thigh, shank, foot) (Cain et al., 2016; Mihy et al., 2026). Sensor data were also oriented to a world reference frame using a manufacturer-provided sensor fusion algorithm (APDM/Clario, Philadelphia, PA).

The four walking bouts used for analysis were identified using trigger button pushes. Gait events were identified by passing the foot world-oriented vertical acceleration data through a one-dimensional continuous wavelet transform algorithm (Boyer and Hafer, 2019). The resulting wavelet peaks above a threshold (half the median magnitude of the peaks of the filtered signal) were defined as gait events. We used a ZUPT (Zero velocity UPdaTe) approach (Rebula et al., 2013) to identify periods of foot flat (i.e., zero foot velocity). We then integrated the foot world frame acceleration signal between periods of foot flat to calculate foot trajectories. Stride lengths, velocities, and directions were determined from foot trajectories between consecutive heel strikes. Steady-state strides were defined as those with an inter-stride difference in stride length (m) and stride velocity (m/s) of less than 0.1. See Supplement of Mihy et al. 2026 for more walking identification details.

Segment excursions were calculated as the stride-by-stride integral of the angular velocity about each segment’s functional mediolateral axis. Joint excursions were calculated for each stride as the difference between the segment excursions about that joint. The knee was calculated as thigh minus shank and the ankle was foot minus shank. Joint ROM were calculated as the maximum-minimum joint excursion. Early stance knee ROM was calculated as the maximum-minimum joint excursion from the first 30% of stance phase.

### 2.4. Statistics

Outcome variables of interest (hip, knee, and ankle ROM, early stance knee ROM, gait speed, and stride length) were calculated per stride and averaged over all steady-state strides for each Walk (1, 2, 3, and 4) for each participant. These variables were selected because they have associations with pain (Bindawas, 2016; Boyer and Hafer, 2019; Maly et al., 2008) and are easy to collect across many settings using IMUs. To determine the change in pain and walking mechanics in response to bouts of stair activity, the change in each variable was calculated as bout1Δ = Walk 2 - Walk 1 and bout2Δ = Walk 4 - Walk 3 resulting in two change values per outcome variable per participant.

To determine if pain and gait changed in response to multiple bouts of stair activity, a 2×2 mixed model ANOVA (group x bout) was run using JMP v18.2.2. If a significant interaction was found, Tukey HSD pairwise comparisons were run. Independent t-tests were used to compare the KOOS and performance on the sit-to-stand and 40m walk clinical tests between groups. For all comparisons, α = 0.05.

Based on previous work and because pain experience is somewhat unpredictable, we planned additional alternative analyses using the same statistical tests as above but examining only the knee OA group, split into those who did and did not experience a change in pain. We defined the pain group as participants who had an increase or decrease in pain of at least 1 for either bout of stairs. Previous work using a 20-minute bout of treadmill walking induced pain in only ∼40% of those with knee OA (Boyer and Hafer, 2019). We expected a higher percentage of our knee OA group to experience pain as stairs are often reported as more painful than walking even early in disease progression (Hensor et al., 2015). Additionally, activity can both increase and decrease pain which may cause conflicting changes in biomechanics. Therefore, we ran addition analyses using the absolute change in biomechanics (i.e., |bout1Δ| = |Walk 2 minus Walk 1| and |bout2Δ| = |Walk 4 minus Walk 3|).

## 3. Results

Nineteen healthy older adults (9 female, 63.9±3.6 years) and 19 adults with self-reported physician-diagnosed symptomatic knee osteoarthritis (13 female, 62.6±4.3 years) participated in this study. See Supplement Table 1 for table of values for all outcome variables of interest. Five healthy (26%) and seven knee OA adults (39%) reported a change in pain in response to at least one bout of stairs (Table 1). The change in pain experienced during walking ranged from -1 to +2 for the healthy group and -3 to +3 for the knee OA group (Figure 2). Performance on the functional tasks can be found in Table 1.

**Table 1.** Healthy vs knee OA demographics.

|  | <u>Healthy</u> | <u>Knee Osteoarthritis</u> |  |
| --- | --- | --- | --- |
| | | ΔPain (N=7) | No ΔPain (N=12) |
| **Sex** | 10M / 9F | 1M / 6F | 5M / 7F |
| **Age (yrs)** | 60.8 (4.7) | 65.1 (3.1) | 62.6 (4.2) |
| **BMI (kg/m<sup>2</sup>)** | 28.9 (3.7) | 26.9 (4.7) | 29.0 (4.5) |
| **KOOS** | 87.5 (15.2) | 67.7 (13.7) | 70.7 (15.0) |
| Pain | 89.8 (12.1) | 66.7 (15.3) | 75.5 (14.2) |
| Symptom | 85.1 (17.8) | 60.7 (17.1) | 72.0 (16.5) |
| ADL | 93.1 (10.7) | 79.8 (17.1) | 81.6 (18.0) |
| QoL | 81.9 (22.8) | 63.4 (9.8) | 56.3 (13.1) |
| **Sit-to-stand (#)** | 13.3 (2.4) | 13.6 (3.0) | 12.1 (3.1) |
| **40-m walk (s)** | 19.2 (2.3) | 21.3 (1.9) | 20.0 (2.4) |

Demographic, questionnaire scoring, and functional test performance differences between groups. MEAN (SD). KOOS – Knee Osteoarthritis and Injury Outcome Score, ADL – Activities of Daily Living, QoL – Quality of Life.

**Figure 2.**
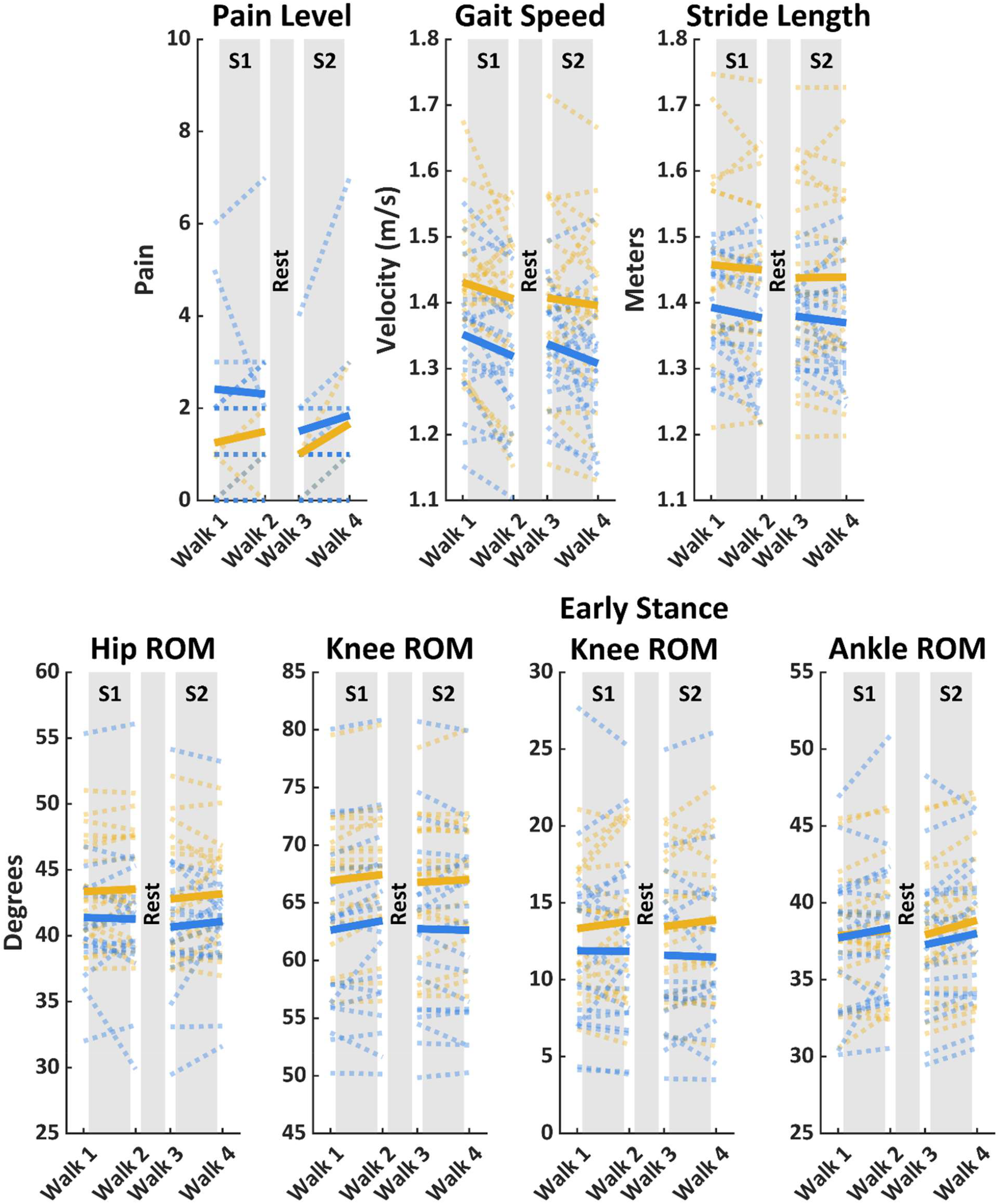
Pain, spatiotemporal variables, and joint ranges of motion (ROM) for healthy adults (yellow lines) and adults with knee osteoarthritis (blue) during the four overground walks. Solid lines represent group averages and dashed lines show each participant’s values. S1 = Stair 1 & S2 = Stair 2.

### 3.1. Healthy vs Knee Osteoarthritis

In our primary analyses, there were no significant interactions of group and bout for any variable of interest (all p ≥ 0.38) (Figure 2, Table 2). Across groups, there was a larger increase in knee ROM in response to the first bout of stairs compared to the second bout of stairs (bout1Δ = 0.7 ± 1.3°, bout2Δ = 0.1 ± 1.3°, p = 0.04). Across bouts, the healthy group had greater increases in early stance knee ROM compared to the knee OA group (healthy = 0.5 ± 0.9°, knee OA = -0.1 ± 1.5°, p = 0.03). When comparing the absolute changes in biomechanics, there were also no significant interactions of group and bout for any variable of interest (all p > 0.10). Across bouts, the knee OA group had a greater absolute change in knee ROM compared to the knee OA group (healthy = 0.8 ± 0.7°, knee OA = 1.2 ± 1.1°, p = 0.03).

**Table 2.**
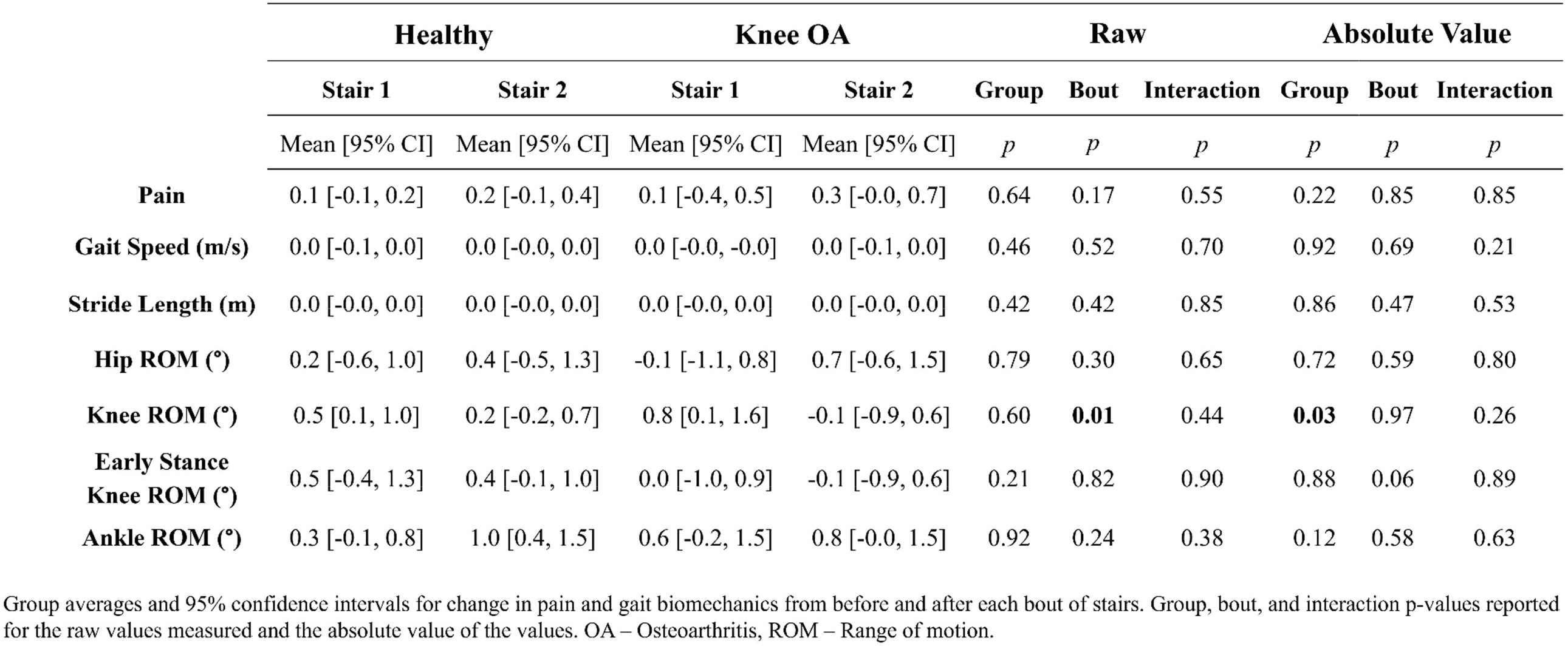
Changes in pain and gait biomechanics for each bout of stairs: Healthy vs Knee Osteoarthritis.

### 3.2. Knee Osteoarthritis Pain vs No Pain

Individuals in the knee OA group were stratified by whether they experienced a change in pain (increase or decrease ≥1) in response to either bout of stairs (pain n = 7; no pain n = 12) (Figure 3, Table 3). There were no significant interactions of group (knee OA pain vs. no pain) and bout for any variable of interest (p > 0.1). Across groups, there was a larger increase in knee ROM in response to the first bout of stairs compared to the second bout of stairs (bout1Δ = 0.9 ± 1.6°, bout2Δ = -0.1 ± 1.4°, p = 0.03). Across bouts, the change in pain group had an increase in early stance knee ROM, but the no change in pain group had a decrease (knee OA pain = 1.0 ± 1.6°, no pain = -0.7 ± 1.6°, p = 0.02). When comparing the absolute magnitude of change, there was a significant interaction for knee ROM (p = 0.02). However, there were no significant post-hoc tests for group or bout repetition for knee ROM (p > 0.29).

**Table 3.** Changes in pain and gait biomechanics for each bout of stairs: Knee Osteoarthritis No ΔPain vs ΔPain.

| | No ΔPain | | ΔPain | | Raw | | | Absolute Value | | |
| --- | --- | --- | --- | --- | --- | --- | --- | --- | --- | --- |
|  | Stair 1 | Stair 2 | Stair 1 | Stair 2 | Group | Bout | Interaction | Group | Bout | Interaction |
|  | Mean [95% CI] | Mean [95% CI] | Mean [95% CI] | Mean [95% CI] | *p* | *p* | *p* | *p* | *p* | *p* |
| **Pain** | 0.0 [0.0, 0.0] | 0.0 [0.0, 0.0] | 0.14 [-1.2, 1.5] | 0.86 [-0.1, 1.8] | 0.11 | 0.11 | 0.11 | **<0.001** | 0.77 | 0.77 |
| **Gait Speed (m/s)** | 0.0 [-0.1, -0.0] | 0.0 [-0.1, 0.0] | 0.0 [-0.1, 0.0] | -0.0 [-0.1, 0.0] | 0.32 | 0.42 | 0.91 | 0.36 | 0.68 | 0.18 |
| **Stride Length (m)** | 0.0 [-0.0, 0.0] | 0.0 [-0.1, 0.0] | 0.0 [-0.1, 0.0] | -0.0 [-0.0, 0.0] | 0.26 | 0.30 | 0.46 | 0.50 | 0.30 | 0.15 |
| **Hip ROM (°)** | -0.1 [-1.6, 1.4] | 0.8 [-0.9, 2.4] | 0.2 [-1.4, 1.0] | 0.4 [-1.1, 1.0] | 0.29 | 0.36 | 0.45 | 0.23 | 0.88 | 0.93 |
| **Knee ROM (°)** | 0.6 [-0.1, 1.3] | -0.1 [-1.3, 1.1] | 1.3 [-0.8, 3.4] | -0.1 [-1.0, 0.7] | 0.35 | **0.03** | 0.52 | 0.74 | 0.29 | **0.02** |
| **Early Stance**<br>**Knee ROM (°)** | -0.9 [-0.3, 1.1] | -0.4 [-0.2, 1.8] | 1.5 [-1.5, 3.6] | 0.3 [-0.7, 2.0] | **0.02** | 0.57 | 0.10 | 0.25 | 0.15 | 0.71 |
| **Ankle ROM (°)** | 0.4 [-1.9, 0.1] | 0.8 [-1.4, 0.6] | 1.1 [-0.1, 3.1] | 0.6 [-1.0, 1.6] | 0.85 | 0.69 | 0.20 | 0.12 | 0.68 | 0.32 |

Group averages and 95% confidence intervals for change in pain and gait biomechanics from before and after each bout of stairs. Group, bout, and interaction p-values reported for the raw values measured and the absolute value of the values. ROM – Range of motion.

**Figure 3.**
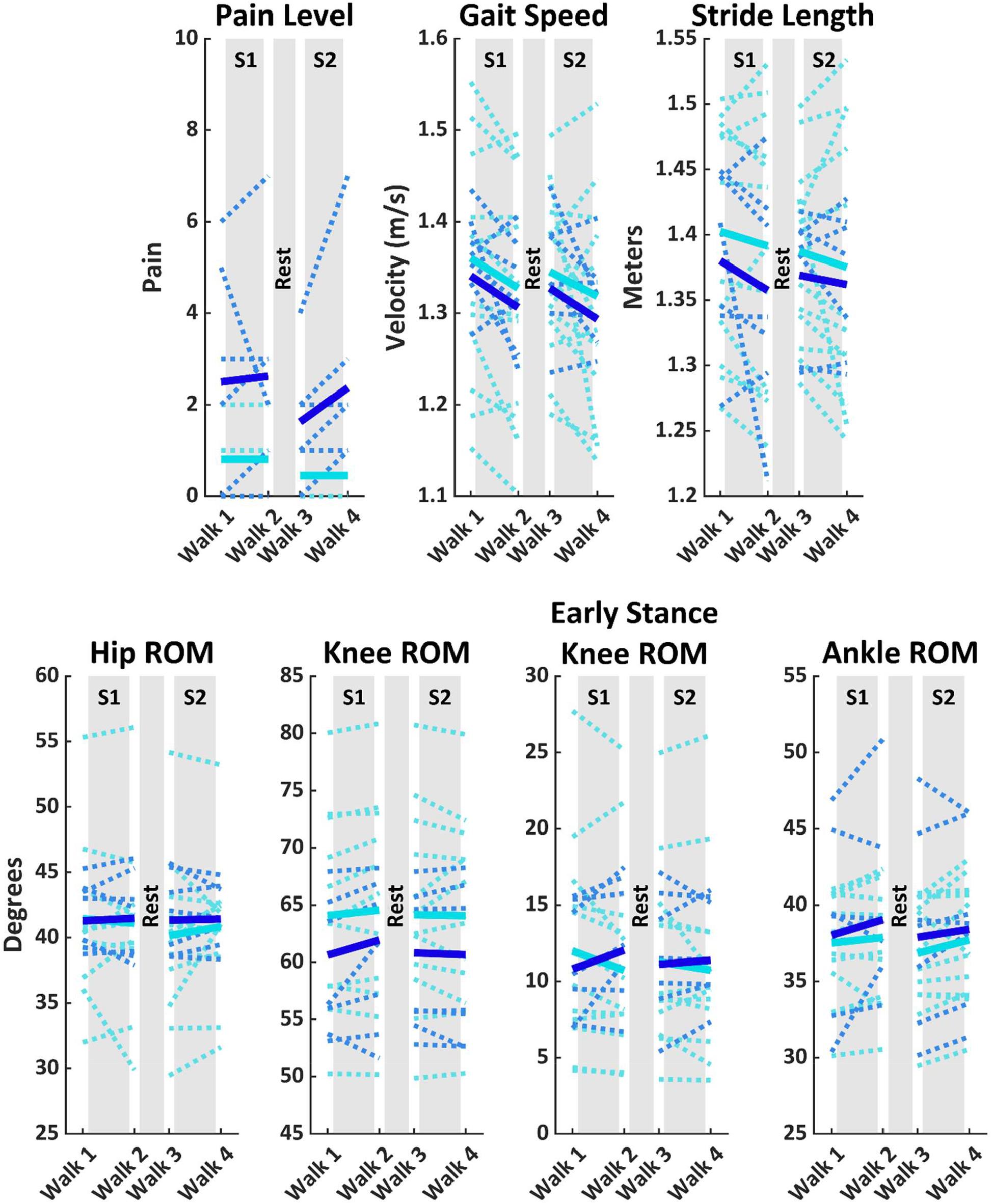
Pain, spatiotemporal variables, and joint ranges of motion (ROM) during the four overground walks for adults with knee osteoarthritis that did (dark blue) and did not (light blue) have a change in pain in response to either bout of stairs. Solid lines represent group averages and dashed lines show each participant’s values. S1 = Stair 1 & S2 = Stair 2.

## 4. Discussion

The primary purpose of this study was to determine how pain and gait change in response to bouts of stair activity in those with and without knee osteoarthritis. There were no differences found in how those with and without knee OA responded to bouts of stairs for pain or walking biomechanics. Across all participants, there was a greater increase in knee range of motion in response to the first bout of stairs compared to the second. Similar to previous pain flare studies, 37% of individuals with knee OA in our study experienced a change in pain in response to activity. When comparing adults with knee OA that did and did not have a change in pain, those who experienced a change in pain had greater changes in early stance knee ROM following stairs compared to those without a change in pain. Those who experienced a change in pain also had a greater absolute change in knee ROM following the first bout of stairs compared to those who did not experience a change in pain. These differences were small (Δ-0.9° to Δ+1.5°) but resemble those found with pain relief (Δ +1°) (Shrader et al., 2004) and with prolonged treadmill walking (Δ < ±1°) (Boyer and Hafer, 2019). These findings suggest that short bouts of stairs can elicit changes in pain similar to extended bouts of treadmill walking and that those with knee OA who experience pain can quickly modify their knee motion during gait.

In contrast to our expectation, we did not find differences in how those with and without knee OA responded to bouts of stairs. The lack of differences between those with and without knee OA may be due to the overall similarity between groups in this study. Functional performance scores were not different between groups, but the knee OA group had expectedly lower KOOS pain scores. Although lower than healthy, these scores were still higher than the values for those with knee OA in other pain flare studies (70.0 for the current study vs. 65.8 knee OA weighted average (Boyer and Hafer, 2019) and 64.4 (Farrokhi et al., 2017) from previous studies). These findings suggest that our knee OA participants had better function and less pain than typically found in the literature which may have impacted our findings.

When splitting the knee OA group into those who did and did not experience a change in pain, we did see a greater absolute change in knee ROM in response to the first bout of stairs in those who experienced a change in pain (+1.3°) compared to those who did not (+0.6°). We also found greater changes in early stance knee ROM following stairs in those that experienced a change in pain (+1.5°) compared to those with no change in pain (-0.9°). Although the between group difference was small, this finding replicates the small differences in knee flexion found by Boyer and Hafer (Δ<1°) (Boyer and Hafer, 2019). Also, in line with their findings, baseline knee ranges of motion for those with knee OA appeared to be lower than the healthy group. Those who did not experience pain were more similar to their healthy counterparts (2-3° less ROM) while those who did experience a change in pain had ∼7° less ROM compared to the healthy group. These results both support previous findings and suggest that IMU measures would be sensitive enough to detect pain-induced changes in less controlled settings. Additionally, those with a change in pain scored 8 points lower for the KOOS pain subsection compared to those whose pain did not change (Table 1). However, the overall KOOS scores were not different between those who did and did not have a change in pain. Therefore, lower KOOS pain scores and baseline differences in knee flexion may be related to the occurrence of pain in response to activity.

In contrast to previous work, we did not control gait speed so that individuals could modify their speed in response to activity or pain. Previous studies either controlled for gait speed or had participants whose gait speeds did not change in response to treadmill walking (Boyer and Hafer, 2019; Farrokhi et al., 2017). Despite not explicitly controlling for speed, our participants did not alter their speed in response to activity, nor did they change their stride lengths. These findings are not what we expected based on studies that relieved pain and found significant changes in gait speed. Our knee OA participants that experienced a change in pain had a -0.04 ± 0.07 m/s change in speed in response to one bout of stairs while previous studies found significant increases in speed that averaged 0.05 m/s and 0.16 m/s in response to pain relief in those with knee OA (Schnitzer et al., 1993; Shrader et al., 2004). Both these pain relief studies and the extended treadmill walk studies had unidirectional changes in pain and biomechanics across their participants (Boyer and Hafer, 2019; Farrokhi et al., 2017; Schnitzer et al., 1993; Shrader et al., 2004). However, this study had bidirectional changes in both pain and biomechanics which may partially explain our lack of expected changes in biomechanics with activity. Therefore, there may be greater variability in the changes in gait speed, stride length, and hip and ankle ROM in response to both pain relief and onset in response to activity compared to knee ROM leading to a lack of significant findings.

Knee joint kinetics (i.e., knee flexion or adduction moments) are the most common measures for determining how gait changes in response to pain. Although we did not measure kinetics, we would expect our findings of increased early stance knee ROM in those who experienced changes in pain to correspond to increases in knee flexion moments (Creaby et al., 2013). These findings contrast those of Boyer and Hafer, where knee OA participants who experienced a change in pain appeared to have a small decrease in stance phase knee ROM and had a significant decrease in knee flexion moment compared to healthy controls in response to a 20-minute treadmill bout (Boyer and Hafer, 2019). Therefore, although both our study and previous studies provide evidence that individuals can quickly modify their gait in the presence of new pain, the direction of change in gait mechanics is inconsistent. Future work should continue to examine the variability in directional changes in joint kinematics, and in turn kinetics, as this variability may explain the inconsistent results in current interventions that aim to provide pain relief and decrease joint loads. Additionally, an understanding of the variability in gait responses to pain will allow for better intervention designs tailored to specific individual needs.

Although the number of participants in our healthy and knee OA groups were similar, there were more females in our knee OA group compared to our healthy group. Our recruitment methods did not differ between groups, but we were unable to recruit sufficient males to have sex matched groups. However, our groups were similar in size to those of Boyer and Hafer (Boyer and Hafer, 2019). Half of the female participants in the knee OA group reported a change in pain in response to activity, but only one of the six males reported a change in pain. These findings are in line with work identifying women report higher pain levels than men regardless of disease severity (Glass et al., 2014), but contrast those of Boyer and Hafer who found fewer women reported a change in pain than didn’t. A limitation of this study was that this study was initially powered for 19 participants per group for our main analysis comparing healthy adults to those with knee OA. However, our secondary analysis comparing those in the knee OA group that did and did not have a change in pain was likely underpowered. Despite being underpowered, our findings resemble the relationships and magnitude of changes found previously in the literature. We also only measured kinematics and spatiotemporal variables to determine whether these IMU-derived metrics were sensitive enough to measure gait changes in response to pain. Although our findings differed from the literature, we were able to demonstrate that IMU-derived measures can detect small changes in response to activity or pain outside of a standard lab setting. Lastly, the KOOS scores in this study indicate our participants were likely in the mild to moderate ranges of knee OA and therefore these findings may not translate to those with more severe knee OA (Kiss, 2011; Zeni and Higginson, 2009).

This study demonstrated that short-duration, high-intensity activities like stair climbing can elicit acute pain changes in individuals with knee osteoarthritis at similar rates to studies that used longer-duration walking to elicit pain. Although we expected stairs to induce greater pain than prolonged treadmill walking, these findings highlight the flexibility to tailor pain protocols to imitate activities of daily living completed by participants (e.g. pain during walking, stairs, standing from chair, etc.) and still observe measurable differences. The observed compensatory changes in knee range of motion despite no changes in spatiotemporal variables suggest that knee kinematics may be more sensitive to acute changes in pain than gait speed or stride length. These compensatory methods were detectable using IMUs and therefore supports the use of IMUs to measure concurrent pain and gait mechanics in less controlled and real-world settings.

## Data Availability

All data produced in the present study are available upon reasonable request to the authors

## Notes

### Competing Interest Statement

The authors have declared no competing interest.

### Funding Statement

This study did not receive any funding

### Author Declarations

IRB of University of Delaware gave ethical approval for this work

